# Genetic polymorphisms mediating behavioural and immune response to pathogens may moderate the impact of the COVID-19 pandemic: a pilot study

**DOI:** 10.1101/2020.06.03.20120998

**Authors:** Ravi Philip Rajkumar

## Abstract

**Background:** The COVID-19 pandemic has affected the entire world, but there are wide variations in prevalence and mortality across nations. Genetic variants which influence behavioural or immune responses to pathogens, selected for by pathogen pressure, may influence this variability. Two relevant polymorphisms in this context are the s allele of the serotonin transporter promoter (5-HTTLPR) and the *G* allele of the interleukin-6 gene (IL-6 rs1800795).

**Methods:** The frequencies of the 5-HTTLPR s allele and IL-6 rs1800795 *G* allele were obtained from published data. The correlations between these allele frequencies and the prevalence and mortality rates of COVID-19 were examined across 44 nations.

**Results:** The IL-6 rs1800795 *G* allele was negatively correlated with COVID-19 prevalence (ρ = −0.466, p < 0.01) and mortality (ρ = −0.591, p<0.001) across nations. The 5-HTTLPR s allele was negatively correlated with COVID-19 mortality rates (ρ = −0.437, p = 0.023).

**Conclusions:** These results suggest that a significant relationship exists between genetic variants that influence behavioural and immune responses to pathogens and indices of the impact of COVID-19 across nations. Further investigation of these variants and their correlates may permit the development of better preventive or therapeutic strategies in the management of the COVID-19 pandemic.

## 1. Introduction

The COVID-19 pandemic has attained the proportions of a global health crisis, with over 6 million people affected to date and over 380,000 deaths reported. Though cases have been reported from virtually every country in the world, the prevalence and fatality rates of this disease have varied significantly across nations [1, 2]. The reasons for this variability are not clear. In the case of variations prevalence, which is directly related to the spread of the disease, explanations related to cultural practices and political will have been proposed [1]. On the other hand, differences in mortality have been attributed to demographic variations or the presence of specific medical comorbidities [3], or to a reduced availability of critical healthcare resources in the face of a sudden surge in cases [1]. However, none of these proposals has been tested at a cross-national level.

A hypothesis that merits further exploration in the context of this pandemic is related to the evolutionary model of population-level differences in allele frequencies. From this point of view, the historical prevalence of infectious pathogens in a specific geographical area has exerted an important influence on genetic variability by influencing the survival of organisms or populations. This may occur if a particular genetic variant affects host immune responses to a pathogen, or even if it favours particular behaviours that potentially minimize the spread of infection across a population. In other words, genetic variants may influence both behavioural and immune responses to the threat of infectious disease [4]. A genetic polymorphism that may influence behavioural patterns of response to pathogens is the s (short) allele of the serotonin transporter promoter region (*5-HTTLPR*). The presence of this allele has been associated with more “collectivist” forms of behaviour and social norms, such as greater obedience to authority and a preference to remain with one’s own group, which may help in curtailing the spread of infection. Its distribution has been found to correlate well with available data on historical pathogen prevalence [5]. From an immunological perspective, the *G* allele of the IL-6 rs1800795 C/G polymorphism, also known as IL-6 -174 C/G, is associated with a more robust immune response to infection, and its prevalence has been found to correlate strongly with both historical and current measures of pathogen load. Though this genetic polymorphism codes primarily for immune responses, it may also influence human cognition and social development through effects on central nervous system inflammation [6].

Given the role that these two genetic variants may have played in protecting populations from outbreaks of infectious disease in the past, this pilot study was conducted to examine their potential impact on two measures of the severity of the COVID-19 pandemic: the prevalence and the crude mortality rate.

## 2. Materials and Methods

To test the association between the 5-HTTLPR s and IL-6 rs1800795 G alleles and the severity of the COVID-19 pandemic, a preliminary analysis of the correlation between the frequencies of these alleles and the prevalence and crude mortality rate reported for COVID-19 across nations was carried out using population-based data. IL-6 rs1800795 G allele frequencies for 35 countries were obtained from the Allele Frequency Net Database, which is a public repository providing information primarily on genes related to immune function [7]. Where more than one estimate of allele frequency for a given nation was available, the weighted mean frequency was computed. Information on 5-HTTLPR s allele frequencies was obtained from the original publication examining this association, which provided aggregate data for 27 countries [5]. Data on at least one of the two allele frequencies was available for 44 countries, and both allele frequencies were available for 18 countries.

Two measures of the impact of COVID-19 were examined: (a) the prevalence, defined as the number of confirmed cases per 1 million population, and (b) the crude mortality rate, defined as the number of deaths caused by COVID-19 per million population. Data on these two variables, for all the countries for which at least one allele frequency was available, was obtained on May 22, 2020 from the Johns Hopkins University of Medicine Coronavirus Resource Center [8], which provides live statistics on the numbers of total cases and deaths due to COVID-19 for all countries. This tracking resource is accessible online at https://coronavirus.jhu.edu/map.html.

Data was entered and analyzed using the Statistical Package for Social Sciences, version 20.0 (IBM SPSS Statistics for Windows, Version 20.0, Armonk, NY: IBM Corporation). All data were tested for a normal distribution using the Kolomogorov-Smirnov test. As none of the variables studied were normally distributed, Spearman’s rank correlation coefficient (ρ) was used to evaluate the association between allele frequencies and COVID-19 prevalence and mortality rates. All tests were two-tailed, and a value of p < 0.05 was considered significant. As a secondary measure, we also examined the correlation between the frequencies of the two genetic variants of interest in this study.

The analysis found that there was a significant negative correlation between the frequency of the IL-6 rs1800785 G allele and both COVID-19 prevalence (ρ = −0.466, p = 0.005, n = 35) and crude mortality rate (ρ = −0.591, p < 0.001, n = 35). Similarly, there was a significant negative correlation between 5-HTTLPR s allele frequency and the crude mortality rate due to COVID-19 (ρ = −0.437, p = 0.023, n = 27). The frequencies of these two alleles were also significantly correlated with each other (ρ = 0.907, p < 0.001, n= 18), suggesting that their distribution may have been subject to common selection pressures - in other words, that behavioural and immune mechanisms to resist pathogen pressure may have co-evolved.

These results, though preliminary, provide confirmation of the hypothesis that population-level variations in allele frequencies, particularly those influence host immune response or favour behaviours that minimize disease spread, may directly influence the spread and severity of the COVID-19 pandemic across nations. These results are preliminary, and point to the need for further study of the interplay between genetics, immunity and behaviour pertaining to infectious pathogens across different populations. A closer study of the mechanisms underlying these effects could lead to the development of optimal behavioural and biological strategies to contain this global pandemic.Materials and Methods should be described with sufficient details to allow others to replicate and build on published results. Please note that publication of your manuscript implicates that you must make all materials, data, computer code, and protocols associated with the publication available to readers. Please disclose at the submission stage any restrictions on the availability of materials or information. New methods and protocols should be described in detail while well-established methods can be briefly described and appropriately cited.

## 3. Results

### 3.1. Relationship between allele frequencies and the prevalence and mortality rates of COVID-19

In the primary analysis, it was found that there was a significant negative correlation between the frequency of the IL-6 rs1800795 G allele and both COVID-19 prevalence (ρ = −0.466, p = 0.005, n = 35) and crude mortality rate (ρ = −0.591, p < 0.001, n = 35). Similarly, there was a significant negative correlation between 5-HTTLPR s allele frequency and the crude mortality rate due to COVID-19 (ρ = −0.437, p = 0.023, n = 27), but not the prevalence. This indicates a potential protective or moderating effect of both these gene polymorphisms on the COVID-19 pandemic, which was more pronounced in the case of the IL-6 rs1800795 *G* allele.

### 3.2. Relationship between 5-HTTLPR s allele and IL-6 rs1800795 G allele frequencies

In the secondary analysis, a strong positive correlation was found between the presence of the 5-HTTLPR s allele and the IL-6 rs1800795 *G* allele (ρ = 0.907, p < 0.001, n= 18). As these genes are on different chromosomes (5-HTTLPR on chromosome 17, IL-6 on chromosome 7), the possibility of genetic linkage was not considered. Instead, this result suggests that these two variants may have been subject to a common selection pressure, in the form of the burden of infectious pathogens.

## 4. Discussion

These results provide support for the contention that genetic variants involved in defence against infectious pathogens – either behaviourally or immunologically – continue to play a significant role in moderating the impact of infectious disease outbreaks in modern times, and more specifically in reducing the spread of, and mortality due to, the novel coronavirus disease COVID-19. It is significant that a more significant relationship was found for the IL-6 rs1800795 *G* allele, which directly modulates the host’s immune and inflammatory response to pathogens, and may thereby influence both the occurrence and the severity of disease following contact with the novel coronavirus SARS-CoV-2. On the other hand, the 5-HTTLPR s allele was only associated with reduced mortality rates. This is a logical consequence of the fact that this gene does not directly affect host immune responses; rather, it may possible influence the likelihood of compliance with measures such as social distancing or isolation, which may serve to contain the spread of the disease to those at a higher risk of poor outcomes, such as the elderly, once the population is aware of the risk of infection. These explanations are provisional and subject to modification based on further data and prospective research.

The results presented above are subject to certain limitations. First, they are based on a hypothesis which, though supported by historical evidence, still remains provisional. Second, they are based on available data from a limited number of countries. Third, as the COVID-19 outbreak is still evolving in many countries, the prevalence and mortality rates used for analysis in this study may not accurately reflect further trends in certain countries, particularly if healthcare systems are overwhelmed by a high case load [1]. Fourth, given the preliminary nature of this study, confounding factors such as mean age and total population, which could influence the prevalence and mortality rates of COVID-19, were not taken into account.

Despite these considerations, this study suggests that further research into the relationship between population-level genetic variants, particularly those related to protection against pathogens, and the impact of disease outbreaks such as the COVID-19 pandemic is warranted. The identification of other genetic polymorphisms influencing either immune or behavioural defences against infection, and the elucidation of their molecular and psychological mechanisms and the interplay between them, could potentially lead to promising new approaches to the control and management of large-scale disease outbreaks, both now and in the future.

## Data Availability

Data will be made available on request.

## Author Contributions

Conceptualization, methodology, data analysis, writing, editing: R.P.R.

## Acknowledgments

In this section you can acknowledge any support given which is not covered by the author contribution or funding sections. This may include administrative and technical support, or donations in kind (e.g., materials used for experiments).

